# Changing molecular epidemiology and characteristics of methicillin-resistant *Staphylococcus aureus* isolated from bloodstream infections: nationwide surveillance in Japan in 2019

**DOI:** 10.1101/2021.11.05.21265983

**Authors:** Norihito Kaku, Daisuke Sasaki, Kenji Ota, Taiga Miyazaki, Katsunori Yanagihara

## Abstract

**Objectives:** Some single-centre studies have reported that MRSA carrying the staphylococcal cassette chromosome mec (SCC*mec*) type IV has been increasing in bloodstream infections (BSIs) in Japan. Therefore, we conducted nationwide surveillance for MRSA BSI to investigate the extent of such change across Japan.

**Methods:** We recruited 51 Japanese hospitals from the Japanese Association for Infectious Diseases. MRSA isolates detected in two or more sets of blood cultures were collected between January and September 2019 and subjected to antimicrobial susceptibility testing. Whole-genome sequencing was also performed to determine SCC*mec* typing and multilocus typing and detect drug-resistance and virulence genes.

**Results:** Two hundred seventy MRSA isolates were collected from 45 hospitals. The major combination types were ST8 with SCC*mec* type IV (ST8-IV) (30.7%), ST1-IV (29.6%), ST2725-IV (9.5%), ST764-II (8.1%), and ST5-II (7.8%). However, there were regional differences among the most major types. The most common types in western, eastern, and northern Japan were ST1-IV, ST8-IV, and ST5-II, respectively. ST8-IV, ST1-IV, and ST2725-IV exhibited greater susceptibility to clindamycin and minocycline than ST764-II and ST5-II, but *erm*(A) was detected in 93.8% and 100% of ST1-IV and ST2725-IV, respectively. Based on drug-resistance and virulence genes, characteristics of ST8-IV were different from those of ST1-IV and ST2725-IV. In addition, there were two major ST8-IV types with different characteristics.

**Conclusions:** This study revealed that SCC*mec* type IV replaced SCC*mec* type II in MRSA BSI. In addition, SCC*mec* type IV was divided into several types with different characteristics.

## Introduction

MRSA is an important drug-resistant pathogen in bloodstream infections (BSIs) because *S. aureus*, including MRSA, is the second-most frequently encountered pathogen in both community- and hospital-onset BSI.^1^ In the United States, the percentage of patients with BSIs involving MRSA increased from 22% in 1995 to 57% in 2001,^2^ but the incidence of MRSA BSIs has been decreasing since the middle of the 2000s through various infection control efforts.^3^ However, a new threat involves the spread of community-associated MRSA (CA-MRSA), represented by the USA300 clone. CA-MRSA strains carrying staphylococcal cassette chromosome *mec* (SCC*mec*) types IV or V have different characteristics than healthcare-associated MRSA (HA-MRSA) strains carrying SCC*mec* types I, II, or III. CA-MRSA strains are more susceptible to antimicrobial agents such as fluoroquinolones, macrolides, lincosamides, and aminoglycosides; moreover, they sometimes produce Panton-Valentine leucocidin (PVL), which causes skin and soft-tissue infections and necrotising pneumonia.^4^ In 2009, a mathematical model predicted that CA-MRSA would become the dominant MRSA strain in hospitals and healthcare facilities,^5^ and in fact, there were some reports that CA-MRSA has replaced HA-MRSA.^6–8^ In addition, the clonal diversity of CA-MRSA is increasing, and it is no longer possible to identify CA-MRSA only by SCC*mec* type IV or production of PVL.^9^

In Japan, sequence type (ST) 5 carrying SCC*mec* type II, represented by the New York/Japan clone, has been the most common clone in MRSA BSIs.^10^ However, the proportion of SCC*mec* type II has been decreasing, while that of SCC*mec* type IV has been increasing in MRSA BSIs.^11,12^ On the other hand, most SCC*mec* type IV strains in Japan lack PVL.^11–13^ However, since these data were obtained from a single institution, the molecular epidemiology and characteristics of MRSA isolated from BSIs in Japan are unclear. In this study, we collected MRSA isolates from patients with BSIs throughout Japan in 2019 and performed antimicrobial susceptibility testing and whole-genome sequencing to reveal the characteristics of these MRSA isolates.

## Materials and Methods

### Study design

We recruited participating medical institutions through a mailing list obtained from the Japanese Association for Infectious Diseases in December 2018; 51 Japanese hospitals were recruited. MRSA isolates detected in two or more blood samples obtained at the same time were collected at each hospital between January 22 and September 30, 2019. Only the first isolate per patient was included in this study. No MRSA isolates met the criteria in the six hospitals. Eventually, 274 isolates collected from 45 hospitals were analysed at Nagasaki University Hospital. Four isolates were excluded from the analysis because they were identified as methicillin-susceptible *Staphylococcus aureus* at Nagasaki University Hospital (Supplementary Figure 1A). Finally, the remaining 270 isolates were analysed (Supplementary Figure 1B). They were stored in Microbank (Iwaki & Co., Ltd., Tokyo, Japan) at -80°C and transferred to Nagasaki University Hospital. The following data regarding MRSA isolates were collected from the participating medical institutions: blood culture collection location (outpatient or inpatient) and day following hospitalization on which blood cultures were collected.

### Antimicrobial susceptibility testing

The MICs for ampicillin, oxacillin, cefoxitin, cefazolin, imipenem, meropenem, levofloxacin, erythromycin, clindamycin, minocycline, trimethoprim/sulfamethoxazole, vancomycin, teicoplanin, linezolid, daptomycin, and arbekacin were determined by broth microdilution testing using Dry Plate Eiken (Eiken, Tokyo, Japan) according to the manufacturer’s instructions. MIC_50_ and MIC_90_ values were calculated as previously reported.^14^ Antimicrobial susceptibility was measured according to CLSI guidelines (M100-Ed31) and EUCAST v.11.0.^15,16^

### WGS and molecular characteristics

All procedures were performed according to the manufacturers’ instructions. DNA was extracted from MRSA isolates using a Quick-DNA Fungal/Bacterial Kit (Zymo Research, Irvine, CA, USA). DNA libraries for sequencing were generated using the Invitrogen Collibri ES DNA Library Prep Kit for Illumina (A38607096, ThermoFisher Scientific, Waltham, MA, USA), and sequencing was performed on a MiSeq system (Illumina, San Diego, CA, USA) using MiSeq Reagent kit v. 3 (600 cycles) (Illumina). Genome assemblies with de Bruijn graphs, MLST, and detection of antimicrobial resistance genes in ResFinder were performed using the CLC Genomics Workbench and Microbial Genomics Modules (QIAGEN N. V., Venlo, Netherlands). SCC*mec* types were determined using SCC*mec*Finder v.1.2 (https://cge.cbs.dtu.dk/services/SCCmecFinder/).^17^ The subtypes of SCC*mec* type IV were also detected using SCC*mec*Finder v.1.2. If two or more subtypes were detected, it is classified as non-subtypeable. Virulence genes were detected using VirulenceFinder v.2.0.3 (https://cge.cbs.dtu.dk/services/VirulenceFinder/, software v.2.0.3, database v.2020-05-29).^18,19^ Core genome MLST (cg MLST) and staphylococcal protein A (spa) typing were performed using Ridom SeqSphere+ ver.8.3.1(Ridom GmbH, Münster, Germany) and genome assemblies for cgMLST and spa typing were performed with SKESA ver.2.3.0.^20^ A Minimum Spanning Tree (MST) was created based on MLST, cgMLST, and *S. aureus* Accessory using Ridom SeqSphere+. Samples with more than 10% missing values of the items for distance calculation were excluded in MST.

### Statistical analysis

All statistical analyses were performed using R software (v. 4.0.3; R Foundation for Statistical Computing, Vienna, Austria). Fisher’s exact test with Bonferroni correction was used to compare categorical variables. The statistical significance level was set at P < 0.05. P values are listed in Table S1.

### Ethics

The Ethics Committee approved this study of Nagasaki University Hospital (19012123). MRSA isolates were anonymized and individually numbered when they were isolated from blood cultures. All data and samples were fully anonymized before being sent to Nagasaki University Hospital. The Ethics Committee of Nagasaki University Hospital waived requirements for informed consent.

### Data availability

Raw data were generated at Nagasaki University Hospital. The derived data supporting the findings of this study are presented in this paper. WGS data reported in this study are available in the DDBJ Sequence Read Archive (https://www.ddbj.nig.ac.jp/dra/index-e.html) under accession number DRA013058.

## Results

### Sequence and SCCmec typing

Among 270 BSI isolates of MRSA, the percentages of SCC*mec* types I, II, IV, V, and IX were 1.9%, 18.5%, 77.4%, 1.9%, and 0.4%, respectively (Table S2). The subtypes of SCC*mec* type IV were IVa (118, 56.5%), IVc (11, 5.3%), IVg (4, 1.9%), IVh (4, 1.9%), and non-subtypeable (72, 34.4%). The major combination types in Japan can be seen in Figure 1A. However, there were regional differences among major types. The most common types in western Japan, eastern Japan, and northern Japan were ST1-IV, ST8-IV, ST5-II, and ST764-II, respectively (Figure 1B). The characteristics of the major types are listed in Table 1. The percentages of hospital-acquired BSIs detected from samples obtained 48 h after hospitalization, involving ST1-IV, ST2725-IV, ST8-IV, ST5-II, and ST764 were 62.5%, 61.5%, 60.2%, 71.4%, 72.7%, respectively. There were no significant differences in patient backgrounds among the types (Table S1). The most detected subtype of SCC*mec* IV in ST8-IV, ST1-IV, and ST2725-IV were non-typeable (80.7%), IVa (100%), and IVa (92.3%) (Table 1).

**Figure 1.**
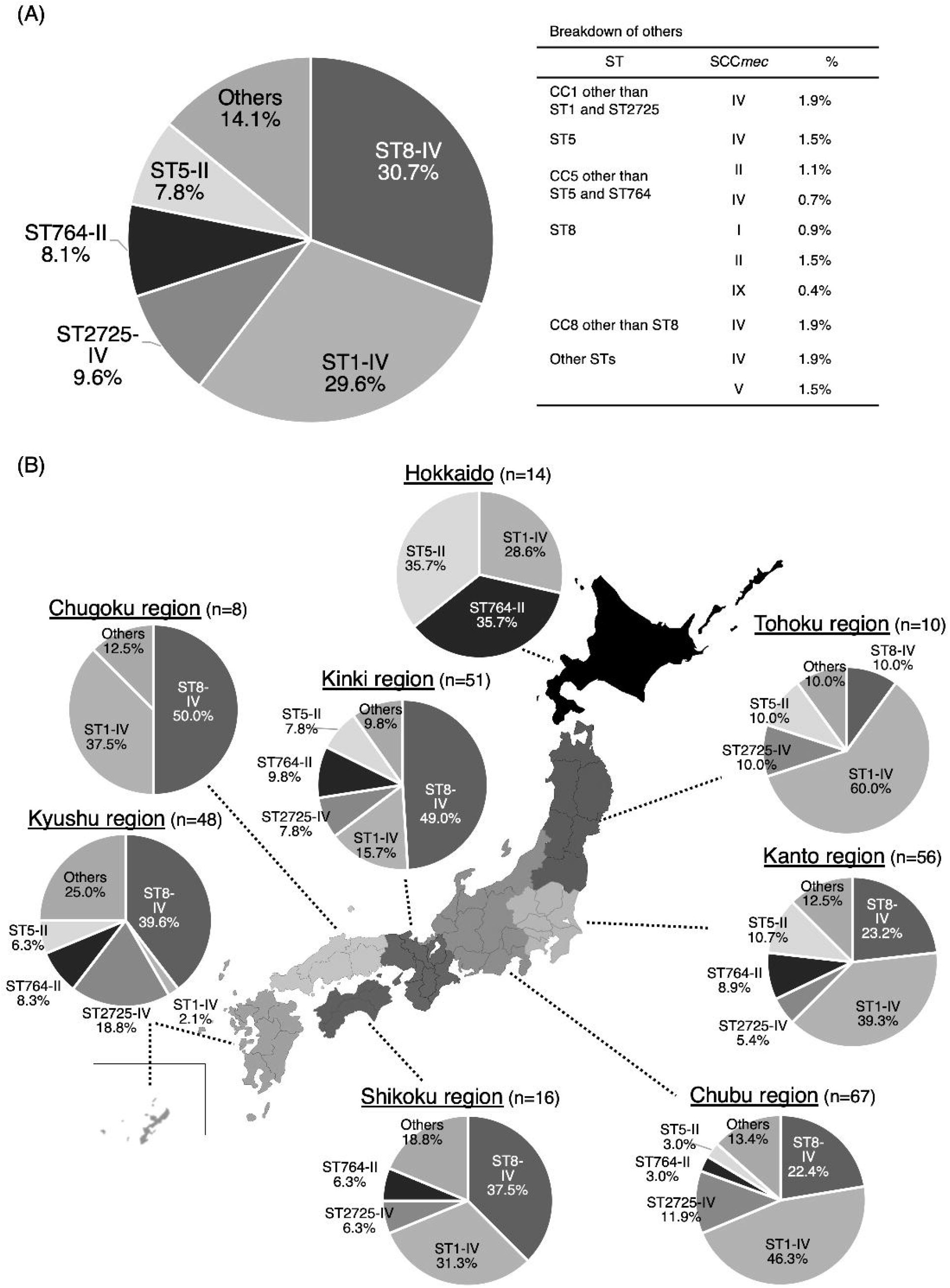
Combination of Sequence type and SCC*mec* type. A percentage of ST and SCC*mec* type in Japan (A) and each region (B). MRSA, methicillin-resistant *Staphylococcus aureus*; ST, sequence type; CC, clonal complex

**Table 1.** Characteristics of the major types.

|  | ST8-IV<br>(n = 83) | ST1-IV<br>(n = 80) | ST2725-IV<br>(n = 26) | ST764-II<br>(n = 22) | ST5-II<br>(n = 21) |
| --- | --- | --- | --- | --- | --- |
| Patient background |  |  |  |  |  |
| Outpatient | 21.7% | 23.8% | 19.2% | 13.6% | - |
| Inpatient within 48 h of hospitalization | 16.9% | 13.8% | 19.2% | 13.6% | 28.6% |
| Inpatient after 48 h of hospitalization | 60.2% | 62.5% | 61.5% | 72.7% | 71.4% |
| Unknown | 1.2% | - | - | - | - |
| Subtypes of SCC<sub>mec</sub> IV |  |  |  |  |  |
| IVa | 8.4% | 100% | 92.3% | - | - |
| IVc | 4.8% | - | - | - | - |
| IVg | 1.2% | - | - | - | - |
| IVh | 4.8% | - | - | - | - |
| Non-subtypeable | 80.7% | - | 7.7% | - | - |
| Spa type |  |  |  |  |  |
| t002 | - | - | - | 59.1% | 61.9% |
| t045 | - | - | - | 4.5% | - |
| t088 | - | - | - | 4.5% | - |
| t458 | - | - | - | - | 4.8% |
| t568 | - | - | - | - | 4.8% |
| t693 | 1.3% | - | - | - | - |
| t1767 | 34.9% | - | - | 4.5% | 4.8% |
| t1779 | 1.2% | - | - | - | - |
| t1784 | 1.2% | 72.5% | 84.6% | - | - |
| t4133 | 2.4% | - | - | - | - |
| t4407 | 1.2% | - | - | - | - |
| t4494 | - | 2.5% | - | - | - |
| t5071 | 24.1% | - | - | - | - |
| t7083 | - | - | - | - | 9.5% |
| t7744 | - | 5.0% | 3.8% | - | - |
| t17749 | 1.2% | - | - | - | - |
| t18300 | 1.2% | - | - | - | - |
| t18648 | 1.2% | - | - | - | - |
| Unknown | 3.6% | 3.8% | - | 4.5% | - |
| Unable to analyze | 15.7% | 15.0% | 11.5% | 22.7% | 14.3% |
| Drug resistance rate in CLSI/EUCAST |  |  |  |  |  |
| Oxacillin | 96.4 / 96.4 | 88.8 / 88.8 | 88.5 / 88.5 | 100 / 100 | 100 / 100 |
| Cefoxitin | 96.4 / 96.4 | 97.5 / 97.5 | 92.3 / 92.3 | 100 / 100 | 100 / 100 |
| Levofloxacin | 55.4 / 56.6 | 100 / 100 | 100 / 100 | 100 / 100 | 100 / 100 |
| Erythromycin | 59.0 / 61.4 | 93.8 / 93.8 | 100 / 100 | 100 / 100 | 100 / 100 |
| Clindamycin | 44.6 / 45.8 | 3.8 / 5.0 | 3.8 / 3.8 | 100 / 100 | 85.7 / 85.7 |
| Minocycline | 0.0 / 30.1 | 0.0 / 6.3 | 0.0 / 3.8 | 4.5 / 100 | 4.8 / 71.4 |
| Trimethoprim /<br>Sulfamethoxazole | 0.0 / 0.0 | 0.0 / 0.0 | 0.0 / 0.0 | 0.0 / 0.0 | 0.0 / 0.0 |
| Vancomycin | 1.2 / 2.4 | 0.0 / 1.3 | 0.0 / 3.8 | 0.0 / 0.0 | 0.0 / 0.0 |
| Teicoplanin | 0.0 / 1.2 | 1.3 / 1.3 | 0.0 / 0.0 | 0.0 / 0.0 | 0.0 / 0.0 |
| Linezolid | 1.2 / 1.2 | 1.3 / 1.3 | 3.8 / 3.8 | 4.5 / 4.5 | 0.0 / 0.0 |
| SCC<sub>mec</sub>, staphylococcal cassette chromosome *mec* |  |  |  |  |  |

#### Spa type and cgMLST

The most common spa type was t784 for ST1-IV and ST2725-IV, and t002 for ST764-II and ST5-II (Table 1). There were two main spa types in ST8-IV: t1767 and t5071. In cgMLST, 203 isolates were classified into 191 complex types. There were nine complex types containing two or three isolates, and isolates of the same complex type were all detected in the same hospital. The number of cgMLST types containing two or more isolates in ST8-IV, ST1-IV, ST2725-IV, ST764-II, and ST5-II was 2, 2, 3, 1, and 1, respectively. MST for all isolates indicated that ST1 and ST2725 were in the same cluster (Figure 2).

**Figure 2.**
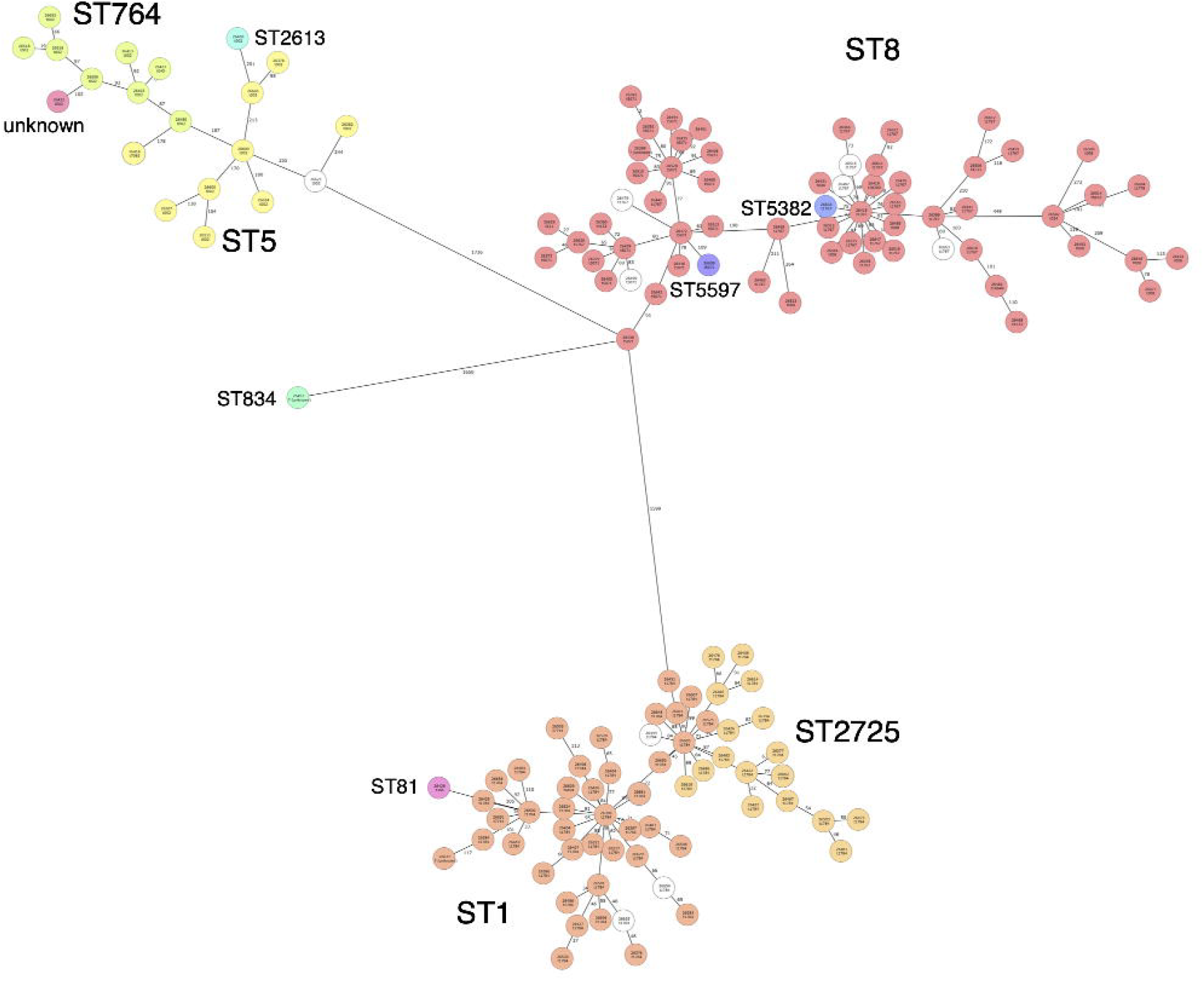
Minimum Spanning Tree for all isolates. One hundred twenty-two samples with more than 10% missing values of the items for distance calculation were excluded from MST. MST for 149 samples was created based on MLST (8), cgMLST (1861), and *S. aureus* Accessory (706) using Ridom SeqSphere+. The number in the node is a complex type determined by cgMLST. MST, Minimum Spanning Tree; cgMLST, core genome MLST.

**Figure 3.**
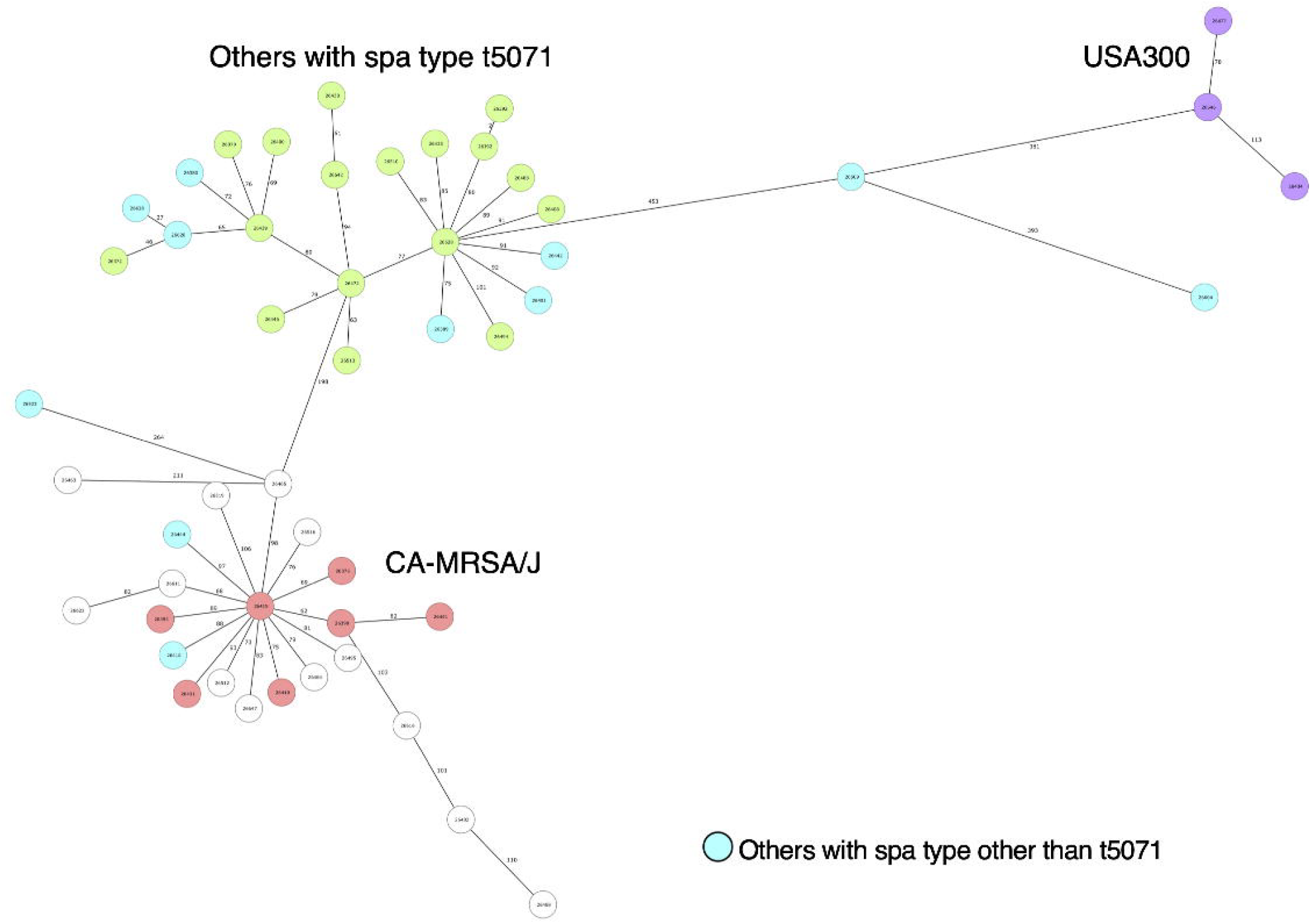
Minimum Spanning Tree for ST8-IV isolates. Twenty-two samples with more than 10% missing values of the items for distance calculation were excluded from MST. MST for 51 samples was created based on MLST (7), cgMLST (1861), and *S. aureus* Accessory (706) using Ridom SeqSphere+. The number in the node is a complex type determined by cgMLST. MST, Minimum Spanning Tree; cgMLST, core genome MLST.

### Antimicrobial susceptibility

Resistance rates calculated using CLSI/EUCAST criteria are shown in Table 1. The resistance rates for anti-MRSA agents were very low in all types. However, for some antimicrobial agents, there were significant differences among the types. The drug resistance rate for levofloxacin in both CLSI and EUCAST criteria in ST8-IV was significantly lower than that of other types (p <0.001). The resistance rates for clindamycin for ST8-IV, ST1-IV, and ST2725-IV were significantly lower than ST5-II and ST764-II (p <0.05). In addition, the drug resistance rate for clindamycin in ST1-IV and ST2725-IV was significantly lower than that for ST8-IV (p <0.001). The resistance rates for minocycline in EUCAST criteria in ST8-IV, ST1-IV, and ST2725-IV were significantly lower than those of ST5-II and ST764-II (p <0.05).

There were differences among the major types in the MICs for β-lactams (Table 2). Both MIC_50_ and MIC_90_ for cefazoline, imipenem, and meropenem were lower in ST8-IV, ST1-IV, and ST2725-IV than in ST5-II and ST764-II. These differences were particularly pronounced for imipenem. The MIC_90_ for imipenem was ≤ 0.25, ≤ 0.25, and 0.5, respectively, in ST8-IV, ST1-IV, and ST2725-IV, respectively, whereas it was ≥ 32 in both ST5-II and ST764-II.

**Table 2.**
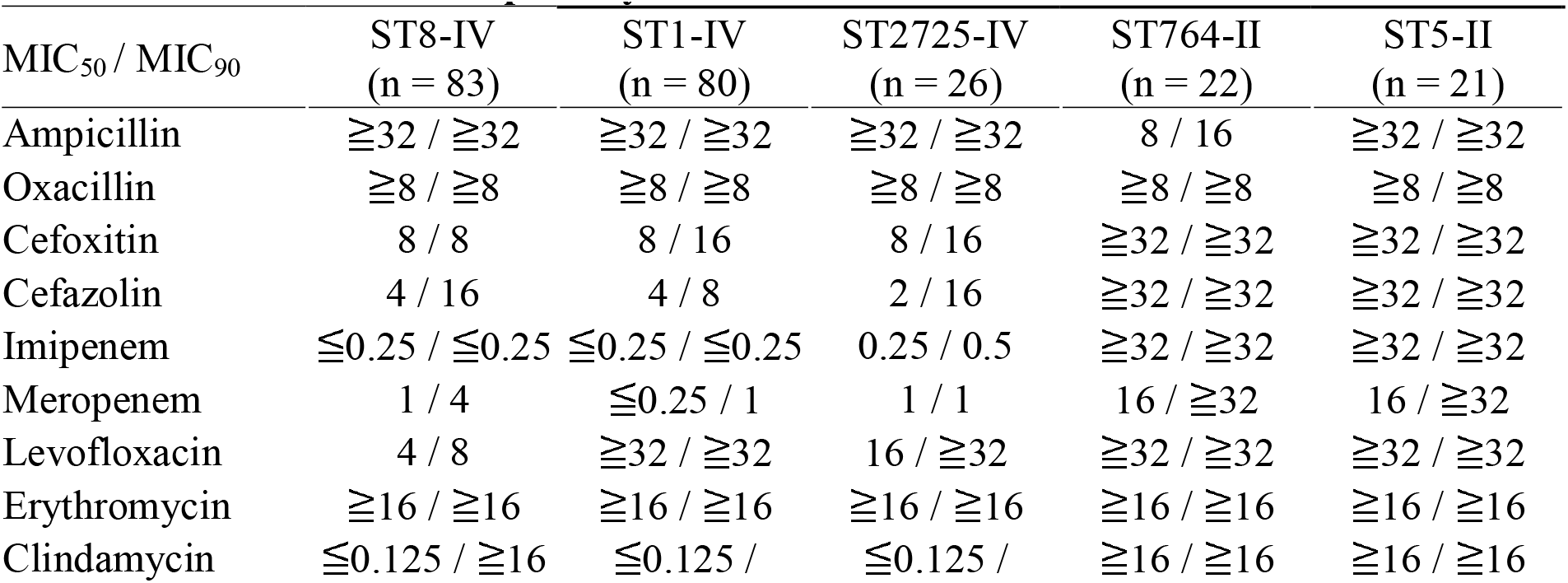
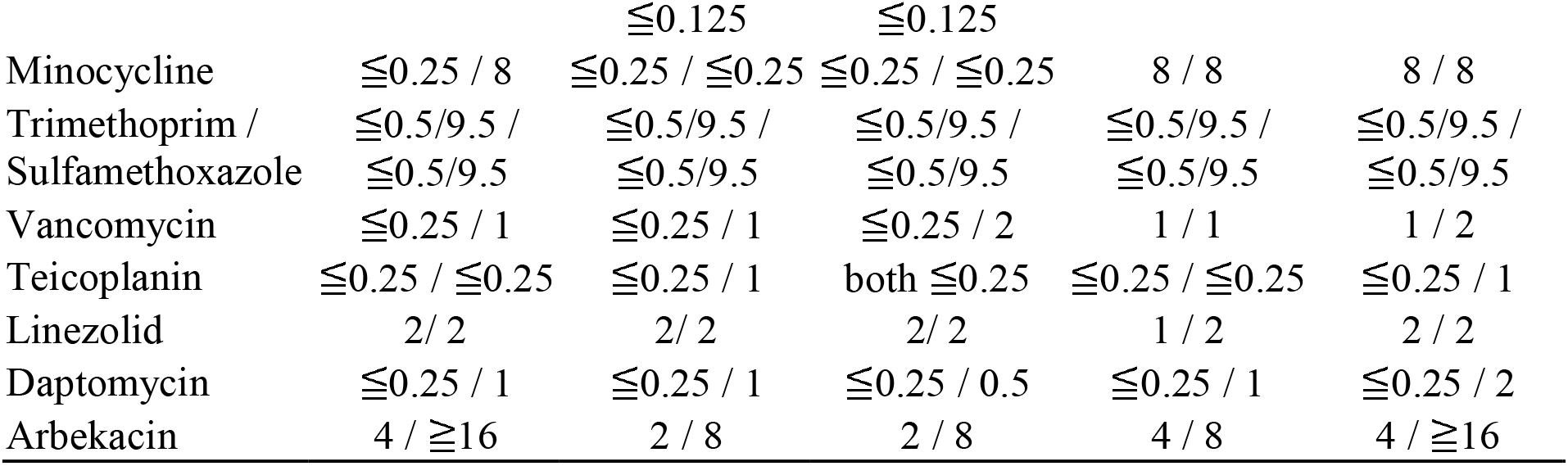
Antimicrobial susceptibility of MRSA isolates.

### Drug-resistance genes

Table 3 shows the positive rate for drug-resistance genes in each type. Almost all major types carried aminoglycoside-resistance genes. However, specific aminoglycoside-resistance genes differed among the major types; for example, the positive rate for *ant(9)-Ia* for ST8-IV was significantly lower than for other types (p <0.001). The positive rate for the macrolide-resistance gene *erm*(A) for ST8-IV was also significantly lower than for other types (p <0.001). In ST764-II, the positive rate for *blaZ* was significantly lower than for other types (p <0.001), whereas that for the fosfomycin-resistance gene was significantly higher than for other types (p <0.05). The positive rate for the tetracycline-resistance gene was significantly higher for ST5-II and ST764-II than for ST8-IV, ST1-IV, and ST2725-IV (p < 0.05).

**Table 3.** The positive rate for drug-resistance genes.

|  | ST8-IV | ST1-IV | ST2725-IV | ST764-II | ST5-II |
| --- | --- | --- | --- | --- | --- |
|  | (n = 83) | (n = 80) | (n = 26) | (n = 22) | (n = 21) |
| Aminoglycoside-resistance gene | 92.8% | 95.0% | 96.2% | 100% | 100% |
| *aac(6')-aph(2'')* | 56.6% | 8.8% | 11.5% | 81.8% | 42.9% |
| *aaD* | 27.7% | 1.3% | - | 40.9% | 85.7% |
| *ant(6)-Ia* | 2.4% | - | - | 4.5% | - |
| *ant(9)-Ia* | 45.8% | 95.0% | 96.2% | 100% | 100% |
| *aph(3')-III* | 2.4% | - | - | - | - |
| β-lactamase gene | | | | | |
| *blaZ* | 97.6% | 98.8% | 92.3% | - | 71.4% |
| Chloramphenicol-resistance gene, |  |  |  |  |  |
| *cat(pC221)* | - | - | - | 4.5% | - |
| Fosfomycin-resistance gene | 6.0% | - | - | 40.9% | 4.8% |
| *fosB4* | - | - | - | 4.5% | 4.8% |
| *fosD* | 6.0% | - | - | 40.9% | - |
| Macrolide-resistance gene | 56.6% | 93.8% | 100% | 100% | 100% |
| *erm(A)* | 47.0% | 93.8% | 96.2% | 100% | 100% |
| *erm(B)* | - | 1.3% | - | 4.5% | - |
| *erm(C)* | 8.4% | 1.3% | 4.4% | 4.5% | - |
| *mph(C)* | 2.4% | - | - | 4.5% | - |
| *msr(A)* | 2.4% | - | - | 4.5% | - |
| Methicillin-resistance gene, *mecA* | 100% | 100% | 100% | 100% | 100% |
| Tetracycline-resistance gene | 28.9% | - | 3.8% | 100% | 76.2% |
| *tet(K)* | 2.4% | - | - | 4.5% | - |
| *tet(M)* | 26.5% | - | 3.8% | 100% | 76.2% |
| *tet(S/M)* | 24.1% | - | - | 100% | 57.1% |
| Trimethoprim-resistance gene, *dfrG* | 1.2% | - | - | - | - |

### Virulence genes

Table 4 shows the positive rate for virulence genes in each type. Almost all isolates carried exoenzyme genes, such as *aur, splA, and splB*. Although > 90% of the isolates in ST1-IV and ST2725-IV also carried *splE*, no isolate in ST5-II and ST764-II carried it. In terms of toxins, almost all isolates carried *hlgA, hlgB, hlgC, LukD*, and *LukE*. Only five (6.0%) isolates in ST8-IV carried *LukF-PV*. CC1-IV, composed of ST1-IV and ST2725-IV, whereas CC5-II, composed of ST5-II and ST764-II, had unique virulence genes. CC1-IV carried *sea, seh, sek*, and *seq*, whereas CC5-II carried *sec, sei, sem, sen, seo*, and *seu*. However, there were some differences between ST5-II and ST764-II; ST5-II carried *sec, sel*, and *tst* more frequently than ST764-II. For other virulence genes, the arginine catabolic mobile element (ACME) was detected in 1.3%, 6.0%, and 31.8% for ST1-IV, ST8-IV, and ST765-II, respectively.

**Table 4.** The positive rate for virulence genes.

|  | ST8-IV<br>(n = 83) | ST1-IV<br>(n = 80) | ST2725-IV<br>(n = 26) | ST764-II<br>(n = 22) | ST5-II<br>(n = 21) |
| --- | --- | --- | --- | --- | --- |
| Exoenzyme genes |  |  |  |  |  |
| *aur* | 100% | 100% | 100% | 100% | 100% |
| *splA* | 96.4% | 98.8% | 96.2% | 100% | 100% |
| *splB* | 94.0% | 98.8% | 96.2% | 100% | 100% |
| *splE* | 47.0% | 91.3% | 96.2% | - | - |
| Toxin genes |  |  |  |  |  |
| *edinA* | 18.1% | - | - | - | - |
| *hlgA* | 98.8% | 100% | 96.2% | 100% | 95.2% |
| *hlgB* | 100% | 100% | 100% | 100% | 100% |
| *hlgC* | 86.7% | 100% | 100% | 100% | 100% |
| *LukD* | 97.6% | 100% | 100% | 100% | 95.2% |
| *LukE* | 98.8% | 100% | 100% | 100% | 95.2% |
| *LukF-PV* | 6.0% | - | - | - | - |
| *sea* | 4.8% | 88.8% | 80.8% | - | 4.8% |
| *seb* | 1.2% | - | 3.8% | 86.4% | 9.5% |
| *sec* | 44.6% | - | 3.8% | 13.6% | 61.9% |
| *seg* | - | - | - | 95.5% | 81.0% |
| *seh* | - | 100% | 96.2% | - | 4.8% |
| *sei* | - | - | - | 95.5% | 100% |
| *sek* | 6.0% | 85.0% | 80.8% | 13.6% | - |
| *sel* | 39.8% | - | - | 13.6% | 71.4% |
| *sem* | - | - | - | 100% | 100% |
| *sen* | - | - | - | 100% | 90.5% |
| *seo* | - | - | - | 100% | 95.2% |
| *sep* | 67.5% | - | - | 9.1% | 28.6% |
| *seq* | 7.2% | 85.0% | 80.8% | 13.6% | 4.8% |
| *seu* | - | - | - | 100% | 85.7% |
| *tst* | 44.6% | 3.8% | 3.8% | 13.6% | 66.7% |
| Other |  |  |  |  |  |
| ACME | 6.0% | 1.3% | - | 31.8% | - |
| *sak* | 94.0% | 98.8% | 100% | 72.7% | 85.7% |
| *scn* | 96.4% | 98.8% | 100% | 86.4% | 85.7% |

### Diversity in ST8-IV

Based on positive rate for drug-resistance and virulence genes, ST8-IV isolates were classified into three groups: USA300 carrying both *LukF-PV* and ACME (4, 4.8%), *LukF-*PV-negative isolates carrying *sec* and *tst* known as CA-MRSA/J (36, 43.4%), and others (43, 51.8%) (Table 5). Of the isolates classified as others, nineteen isolates were spa type t5071. MST for ST8-IV isolates indicated that others with spa type t5071 were classified to a different cluster than USA300 and CA-MRSA/J. The characteristics of others with spa type t5071 differed from CA-MRSA/J in that they showed a high rates of resistance to levofloxacin (100%), erythromycin (94.7%), clindamycin (76.7%), minocycline (78.9%) in EUCAST criteria. Other isolates of spa type t5071 had higher rates of resistance genes, such as *ant(9)-Ia* (100%), *erm(A)* (100%), *tet(M)* (68.4%), and *tet(S/M)* (63.2%); they also showed the higher positive rate for *splE* (84.2%) than CA-MRSA/J. Of other isolates with spa type t5071, 42.1% were detected in the Kinki region and 73.7% were isolated from in-patients after 48 h of hospitalization, although there were only two isolates of the same complex type in cgMLST.

**Table 5.**
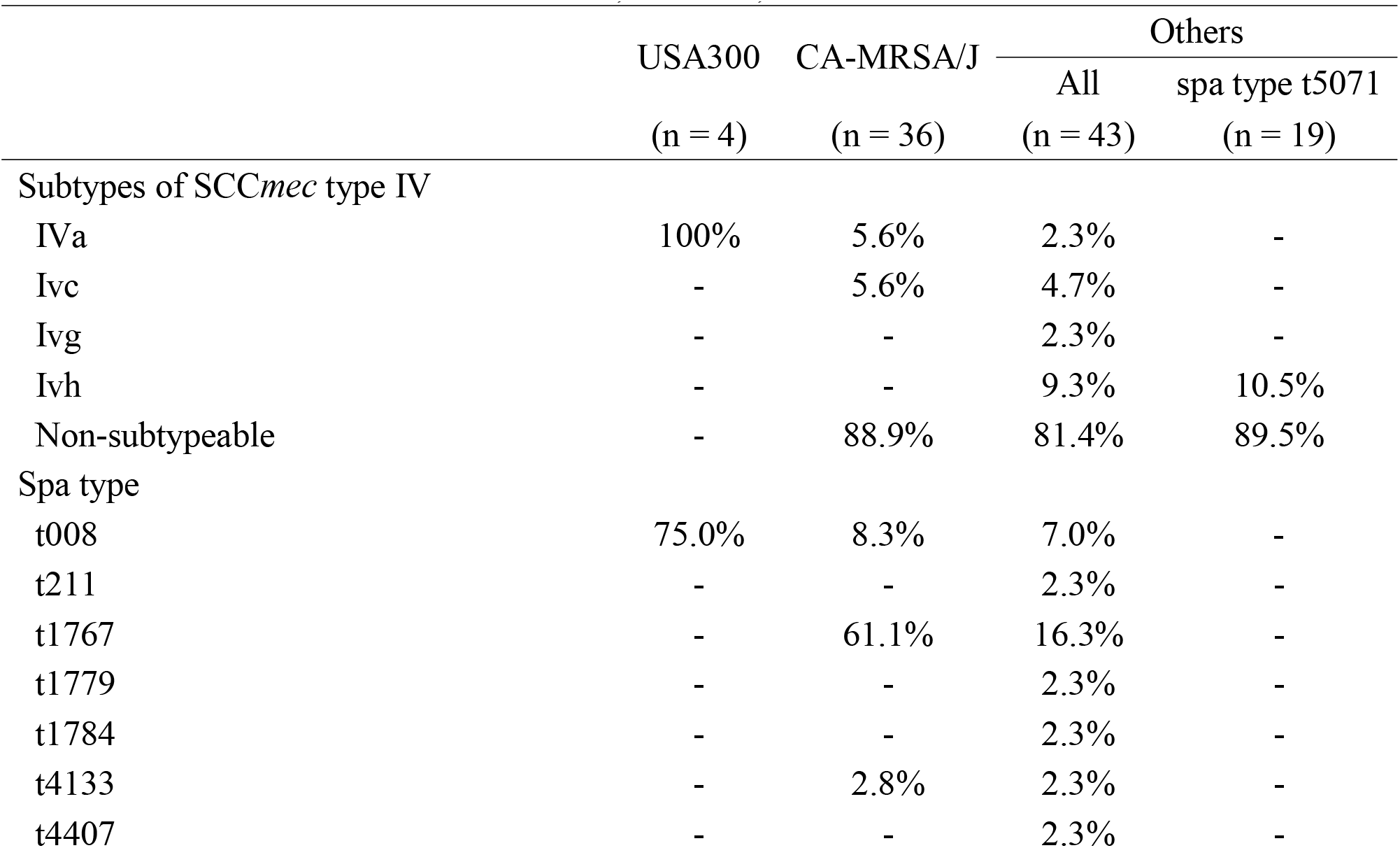
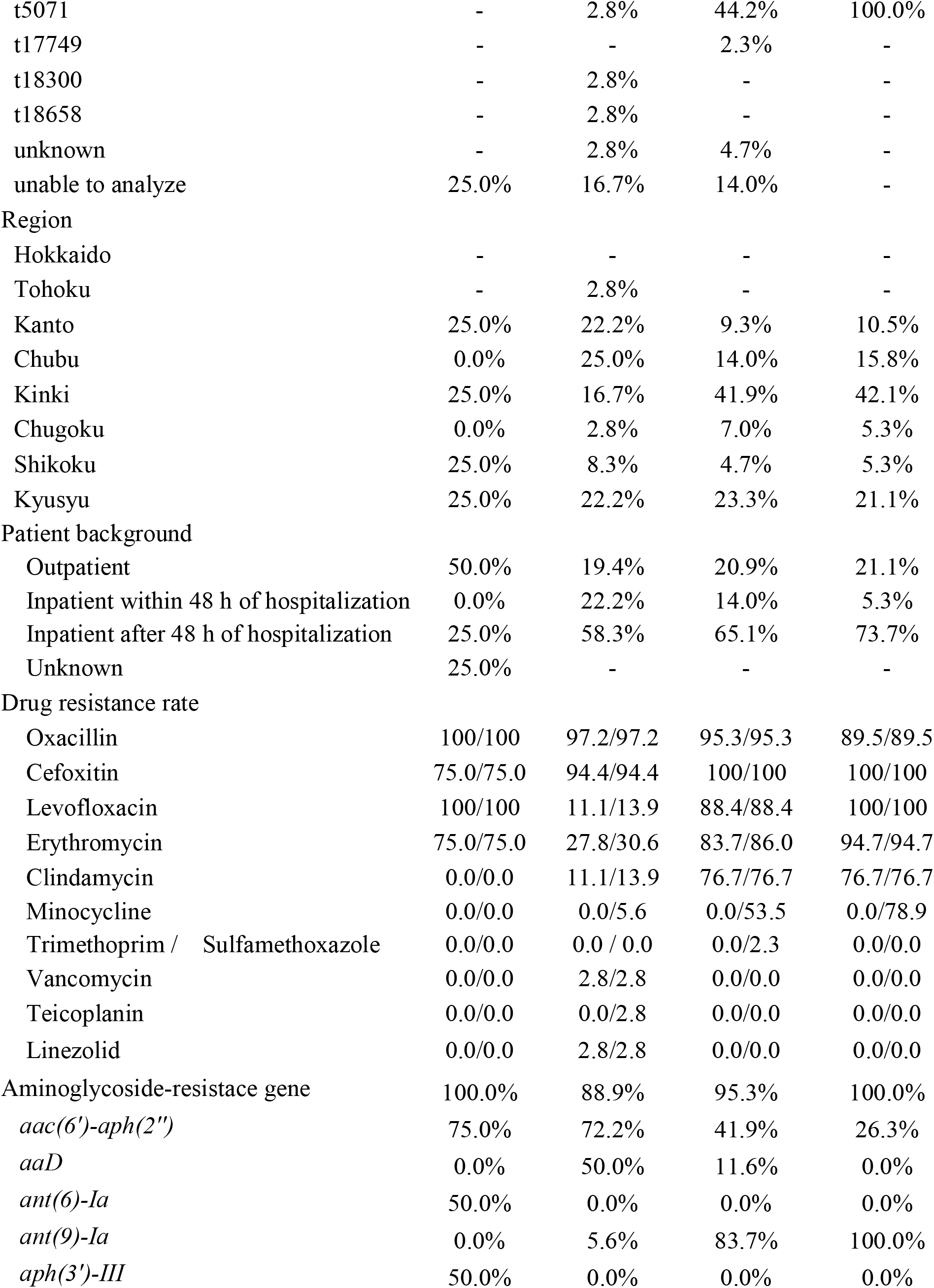
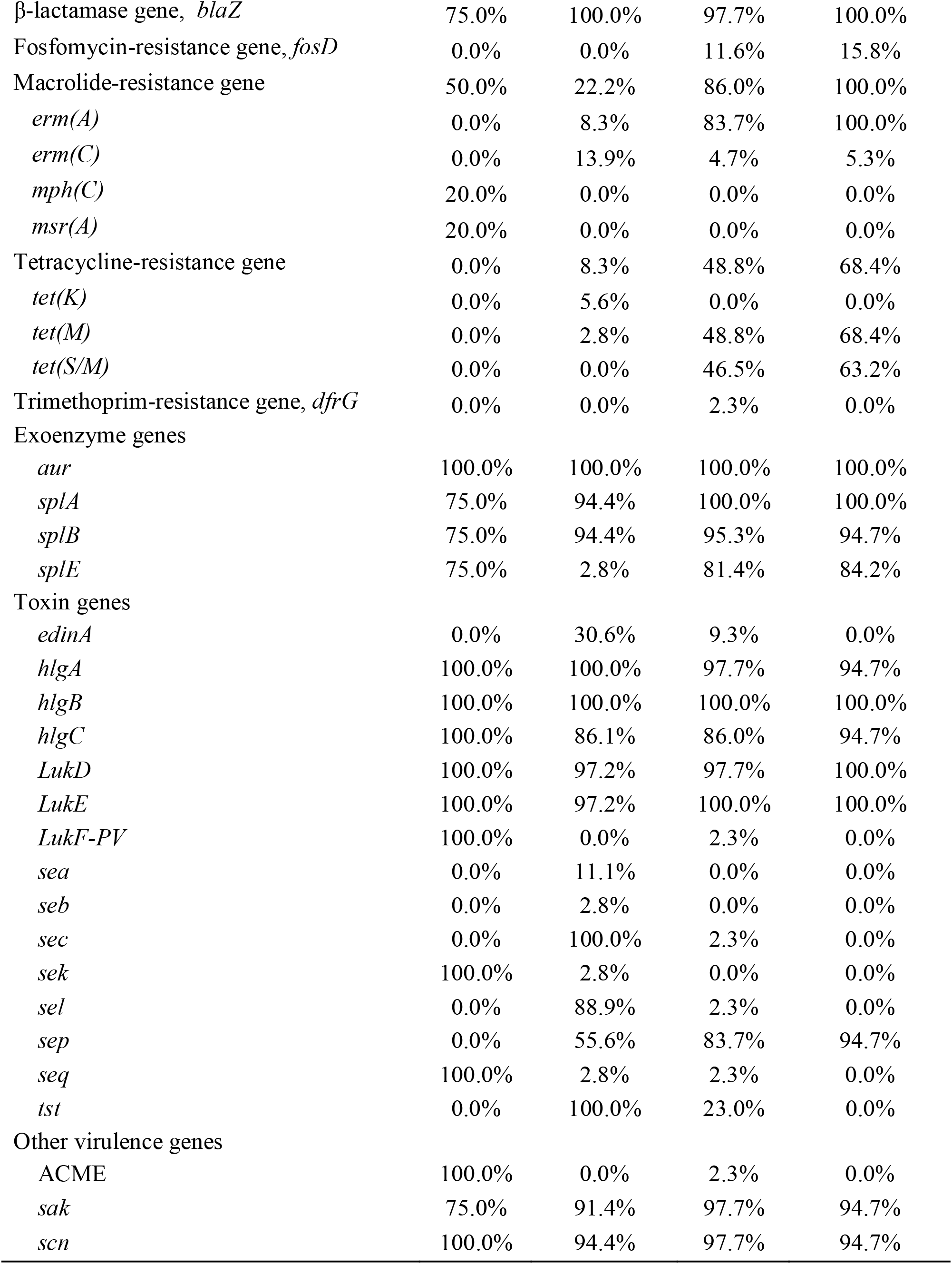
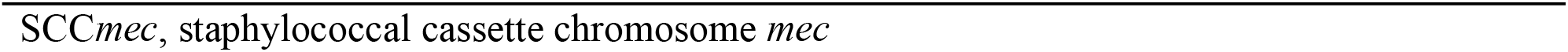
Characteristics of CA-MRSA/J, USA300, and other isolates in ST8-IV.

## Discussion

The most common MRSA type isolated from BSI in Japan in 2019 was ST8-IV, closely followed by ST1-IV. In contrast, ST5-II, which had been the most prevalent in Japan,^10^ ranked fifth, representing only 7.8%. A previous Japanese study, which analysed 151 MRSA isolates detected in blood cultures from 53 medical institutions in 2011, reported that the percentage of SCC*mec* type IV was only 19.9%, whereas that of SCC*mec* type II was 75.6%.^21^ However, a previous study from Tokyo in the Kanto region reported that the relative abundances of SCC*mec* types II and IV had reversed between 2015 and 2017, with SCC*mec* type IV the most prevalent, comprising 73.5% of MRSA BSI cases.^12^ Therefore, although participating medical institutions and analytic methods differed between earlier surveillance programs and this study,^21^ our data indicated that ST8-IV and ST1-IV have replaced ST5-II in MRSA BSIs in Japan. However, there were regional differences among the major types. The most common types in Northern, Eastern, and Western Japan were ST5-II and ST764-II, ST1-IV, and ST8-IV, respectively. The most common type in northern Japan, in the Hokkaido region, was very different from other regions, and the percentage of SCC*mec* type II was 71.4%. This is similar to the surveillance results conducted in the Hokkaido region between 2017 and 2019,^22^ although the number of medical institutions that participated in the Hokkaido region in this study was only two. Thus, there may be regional differences in the spread of ST8-IV and ST1-IV. SCC*mec* type IV has been associated with community-acquired infection,^23^ but in this study, more than 60% of ST8-IV, ST1-IV and ST2725-IV cases were hospital-acquired BSI. Some cgMLST types contained two or more isolates of ST8-IV, ST1-IV, and ST2725-IV, but their numbers were small. Therefore, although there had been cases of SCC*mec* type IV spreading in outbreaks in hospitals, the rate is not considered high. On the other hand, the rate of hospital-acquired BSI in SCC*mec* type IV is about 10% lower than SCC*mec* type II in this study. Considering the very high rate of hospital-acquired BSI (82.6%) in Japan for MRSA,^24^ ST8-IV, ST1-IV and ST2725-IV spread in hospitals and communities.

We observed differences among the major types in terms of drug resistance. In antimicrobial susceptibility testing, SCC*mec* type IV (ST8-IV, ST1-IV, and ST2725-IV) exhibited a significantly lower resistance rate for clindamycin and minocycline than SCC*mec* type II of ST5-II and ST764-II. The resistance rate for clindamycin was almost the same as the *erm*(A) gene retention rate in ST8-IV, whereas the resistance rate for clindamycin was much lower than the *erm(A*) gene retention rate in ST1-IV and ST2725-IV. Since the *erm*(A) gene is related to inducible clindamycin resistance,^25^ these results suggested that D-zone test should be performed to detect inducible clindamycin resistance in areas where ST1-IV and ST2825-IV are spreading. In contrast, the drug-resistance rate for minocycline in EUCAST criteria was almost the same as the positive rate for the tetracycline resistance gene. SCC*mec* type IV also showed lower MIC_50_and MIC_90_ values for cefazolin, imipenem, and meropenem than SCC*mec* type II. This result was similar to a previous study.^21^ Since β-lactams exhibit a synergic effect in combination with anti-MRSA agents in several studies,^26,27^ β-lactams might be considered to treat MRSA BSI caused by SCC*mec* type IV in combination with anti-MRSA agents.

Almost all isolates involving all the major types carried *aur, splA, splB, hlgA, hlgB, hlgC, LukD, LukE, sak*, and *scn*. ST1-IV and ST2725-IV, both of which are CC1-IV, had a similar pattern of virulence genes. CC1-IV carried *splE, sea, seh, sek*, and *seq* more frequently than the other types. In addition, MST for all isolates indicated that ST1 and ST2725 were in the same cluster. Therefore, there might be no need to distinguish between ST1-IV and ST2725-IV in Japan. At the clonal complex level, CC1-IV was the most abundant type (39.3%). However, since no isolate carried *LukF-PV* and only one isolate carried ACME in CC1-IV, they differed from USA400.^28^ Several Japanese studies have reported the same characteristics for CC1-IV.^29,30^ In addition, *LukF-PV*-negative CC1-IV has been reported in Europe and Australia.^31,32^ The characteristics of CC1-IV in this study and previous Japanese studies differ from those of European *LukF-PV*-negative CC1-IV regarding virulence genes,^32^ whereas Japanese CC1-IV have a similar pattern to *LukF-PV*-negative CC1-IV in Australia.^31^ However, there exists a difference in macrolide-resistance genes between Japan and Australia. In Australia, *erm*(C) was detected in CC1-IV,^31^ but *erm*(A) was detected in CC1-IV in this study. These results indicate that CC1-IV has evolved and spread independently in Japan. ST5-II and ST764-II, both of which are CC5-II, also had a similar pattern of virulence genes, but there were significant differences between them. ST5-II carried *sec, sel*, and *tst* more frequently than ST764-II. They also had different drug resistance genes, such as *aaD, fosD, and tet(S*/M). Since the spread of ST764-II among elderly Japanese in long-term care facilities in Japan,^33^ the change in the ratio of CC5-II should be carefully monitored.

ST8-IV was the most frequently detected type in this study, but it carried diverse virulence genes. The positive rate for *sec* and *tst* was 44.6%, with one isolate carrying both *LukF-PV* and ACME. The *LukF-*PV-negative ST8 clone CA-MRSA/J, characterized by carrying *sec* and *tst*, has emerged and spread in Western Japan since 2003;^34^ therefore, it is noteworthy that 43.4% of ST8-IV in this study were estimated as CA-MRSA/J. Four isolates carried both *LukF-PV* and ACME in *sec* and *tst-*negative ST8-IV, and three isolates with spa type identified were all t008.^35^ Based on these results, 4.8% of ST8-IV estimated as USA300. The characteristics of ST8-IV isolates classified as others differed from CA-MRSA/J and USA300 isolates. Focusing on the spa type t5071, the most common spa type in the others, they were classified to a different cluster than USA300 and CA-MRSA/J. The others with spa type t5071 showed higher resistance to several antimicrobial agents, such as fluoroquinolones, macrolides, lincosamides, and aminoglycosides, than CA-MRSA/J. Both CA-MRSA/J and others with spa type t5071 were detected mainly in Western Japan, but others with spa type t5071 were prevalent in the Kinki region. In addition, others with spa type t5071 tended to be detected in inpatients after 48 h of hospitalization. However, there were only two isolates of the same complex type in cgMLST. These findings suggest the spread of two major types of ST8-IV, CA-MRSA/J and spa type t5071 ST8-IV, mainly in Western Japan.

This study had several limitations. First, the impact of changes in SCC*mec* type on the clinical course is unknown because we did not collect the information on patient backgrounds, such as age, underlying diseases, as well as severity and prognosis of BSI. Second, we evaluated the diversity of each MRSA type based on patterns of drug-resistance, virulence genes, spa type, and cgMLST. However, complex type in cgMLST and spa type were not identified in 24.8% and 19.6% of the isolates in Ridom SeqSphere+. We had optimized sequencing in the MiSeq system to determine MLST and detect drug-resistance genes in the CLC workbench. Even though the CLC workbench was able to identify ST of MLST, Ridom SeqSphere+ did not identify ST in 59 (21.9%) of the isolates. The algorithms for genome assembly are different, which may have affected the results. These suggest that the results of WGS may differ depending on the software, algorithm, and database. Therefore, it is necessary to validate the best analysis tool for MRSA WGS in the future. Third, our analyses differed from those used in previous Japanese surveillance programs, as this is the first Japanese surveillance using WGS. In addition, the backgrounds of participating medical institutions in this surveillance are different from those of previous Japanese programs. The participating medical institutions in this study have in-house microbiology laboratories and are considered secondary or tertiary hospitals. On the other hand, the participating medical institutions in the previous study are considered primary hospitals or long-stay hospitals because they outsourced microbiological testing.^21^ It is necessary to continue this surveillance to validate changes in MRSA because the patient populations in the two studies should be different. Fourth, we focused on MRSA BSI. Since BSIs occur secondary to other infections, such as pneumonia and skin and soft-tissue infections, or medical procedure, the major types we found in this study may be spread around medical institutions. Therefore, it is necessary to confirm whether those types are really throughout the community in Japan by conducting nationwide active epidemiological surveillance, including a healthy population.

## Conclusion

This study revealed that SCC*mec* type IV replaced SCC*mec* type II in MRSA BSI. However, the characteristics of the major types in Japanese SCC*mec* type IV were different from USA300 and USA400, with a low prevalence of *LukF-PV* and ACME. This study also indicated that the three main types of SCC*mec* type IV, CC1-IV, CA-MRSA/J, and spa type t5071 ST8-IV, have spread throughout Japan, although there were regional differences.

## Data Availability

The derived data supporting the findings of this study are presented in this paper. WGS data reported in this study are available in the DDBJ Sequence Read Archive (https://www.ddbj.nig.ac.jp/dra/index-e.html) under accession number DRA013058.

## Transparency declarations

The authors have no conflicts of interest directly relevant to the content of this article.

## Acknowledgements

We thank Professor Kazuyoshi Kawakami (Medical Microbiology, Mycology and Immunology, Tohoku University Graduate School of Medicine) for advice on study design and sample collection as chair of the Committee for Clinical Research Promotion of the Japanese Association for Infectious Diseases, Shuji Miyazaki (Department of Laboratory Medicine, Nagasaki University of Hospital) for support of the analysis of bacteria, and Editage (www.editage.com) for English language editing.

We also thank all staff members cooperated in this study at the Japanese Association for Infectious Diseases, Furano Hospital, Sapporo Medical University Hospital, Aomori Prefectural Central Hospital, Odate Municipal General Hospital, Fukushima Medical University, Saitama Medical University International Medical Center, Jikei University Katsusika Medical Center, Yokohama City University Hospital, Yokohama Municipal Citizen’s Hospital, Yokohama City University Medical Center, Tokyo Bay Urayasu Ichikawa Medical Center, Showa University Hospital, Tokyo Saiseikai Central Hospital, Hamamatsu University Hospital, Ishikawa Prefectural Central Hospital, Chuno Kosei Hospital, Asanogawa General Hospital, Nagaoka Red Cross Hospital, University of Fukui Hospital, Toyama University Hospital, Nagoya University Hospital, Nagoya City University West Medical Center, Niigata University Medical & Dental Hospital, Japanese Red Cross Society Suwa Hospital, Daiyukai General Hospital, Osaka Medical and Pharmaceutical University Hospital, Osaka City General Hospital, Kobe City Nishi-Kobe Medical Center, Osaka Univerisity Hospital, Kindai University Hospital, University Hospital Kyoto Prefectural University of Medicine, Nara Prefectural General Medical Center, Shimane University Hospital, Tottori University Hospital, Ehime University Hospital, Chikamori Hospital, Uwajima City Hospital, Oita Prefectural Hospital, Saga-Ken Medical Centre Koseikan, University of The Ryukyus Hospital, Fukuoka University Hospital, National Hospital Organization Nagasaki Medical Center, Fukuoka Children’s Hospital, Nagasaki Goto Chuoh Hospital, and Nagasaki University Hospital for collection of MRSA isolates.

## Contributions

All authors were involved in the study design, acquisition, and interpretation of the data. NK, DS, KO, and KY were involved in data analysis. NK wrote the original manuscript, and all authors revised the manuscript and have approved the manuscript for publication.

## Figure Legends

**Figure 1. The major types in Japan**

The major types in Japan (A) and each region (B) were determined by a combination of sequence type (ST) and SCC*mec* type.

**Figure 2. Minimum Spanning Tree for all isolates**.

One hundred twenty-two samples with more than 10% missing values of the items for distance calculation were excluded from MST. MST for 149 samples was created based on MLST (8), cgMLST (1861), and *S. aureus* Accessory (706) using Ridom SeqSphere+. The number in the node is a complex type determined by cgMLST.

MST, Minimum Spanning Tree; cgMLST, core genome MLST.

**Figure 3. Minimum Spanning Tree for ST8-IV isolates**

Twenty-two samples with more than 10% missing values of the items for distance calculation were excluded from MST. MST for 51 samples was created based on MLST (7), cgMLST (1861), and *S. aureus* Accessory (706) using Ridom SeqSphere+. The number in the node is a complex type determined by cgMLST.

MST, Minimum Spanning Tree; cgMLST, core genome MLST.

**Supplementary Figure 1.**
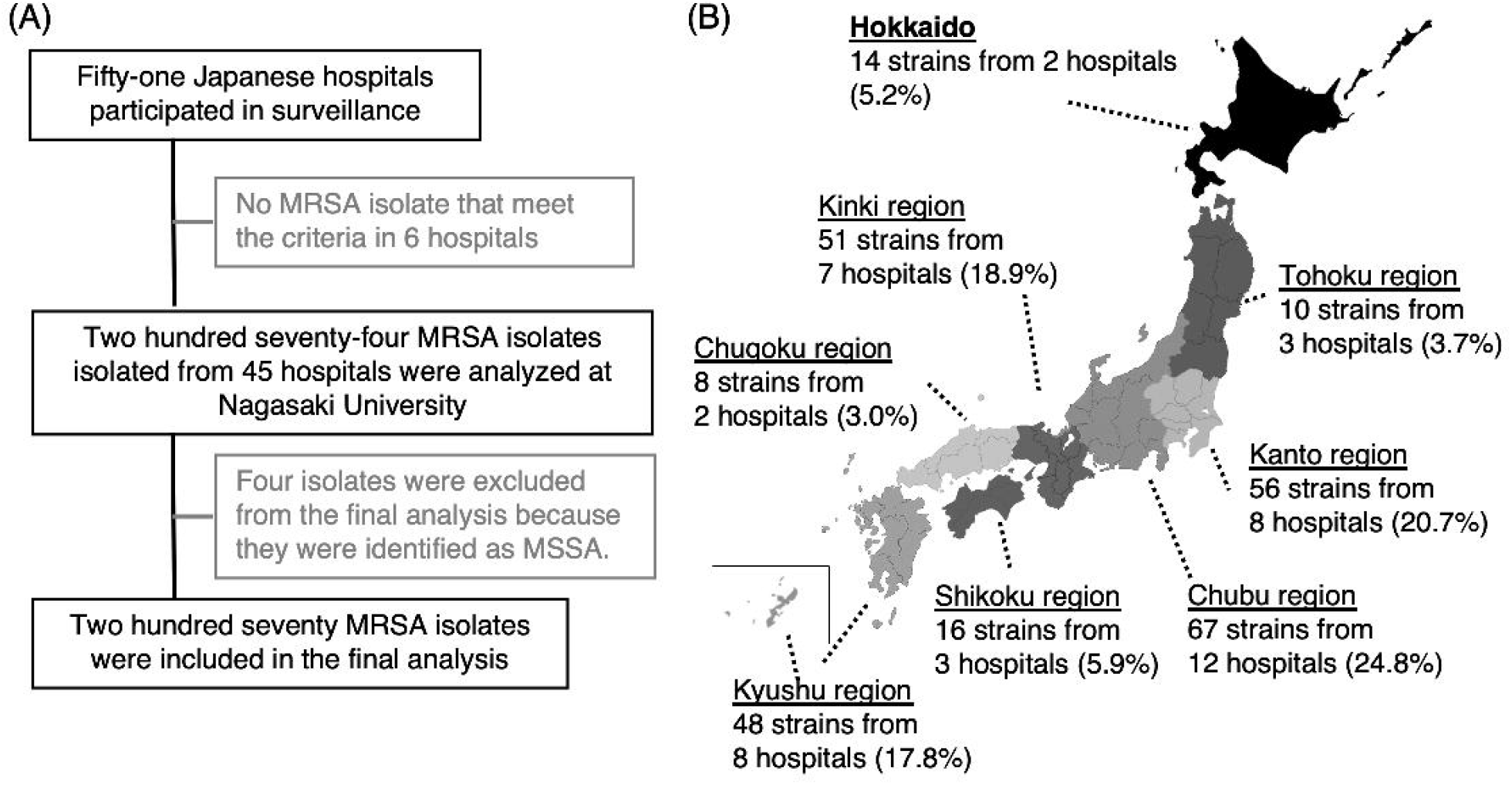
Selection flow diagram and participating hospitals. MRSA, methicillin-resistant *Staphylococcus aureus*; MSSA, methicillin-susceptible *Staphylococcus aureus*

